# A prediction model for COVID-19 prevalence based on demographic and healthcare parameters in Iran

**DOI:** 10.1101/2021.01.27.21250551

**Authors:** Parimah Emadi Safavi, Karim Rahimian, Alireza Doustmohammadi, Mahla Safari dastjerdei, Ahmadreza Rasouli, Javad Zahiri

## Abstract

Coronavirus Disease 2019 (COVID-19) pandemic has become the greatest threat to global health in only a matter of months. Iran struggling with COVID-19 coincidence with Nowruz vacations has led to horrendous consequences for both people and the public health workforce. Modeling approaches have been proved to be highly advantageous in taking appropriate actions in the early stages of the pandemic. To this date, no study has been conducted to model the disease to investigate the disease, especially after travel restrictions in Iran. In this study, we exploited the opportunities that Artificial neural networks offer to investigate contributing factors of early-stage coronavirus spread via generating a model to predict daily confirmed cases in Iran. We collected publicly available data of confirmed cases in 24 provinces from April 4, 2020, to May 2, 2020, with a list of explanatory factors. The factors were checked separately for any linear associations and to train and validate a multilayer perceptron network. The accuracy of the models was evaluated, the R2 scores were 0.842 for population distribution, 0.822 for health index, and 0.864 for the population in the provinces. Our results suggest the significant impact of the mentioned factors on disease spread in the time of travel restrictions when the vacation ended. Accordingly, this information can be implicated in assessing the risk of epidemics and future policy makings in this area.

## Introduction

In late January 2020, Coronavirus disease 2019 (COVID-19) from China became to be an international concern(Anon n.d.). The virus rapidly spread across the world with more than 60 million confirmed cases in 218 countries. Despite all countries’ efforts to mitigate the disease spread through imposing travel restrictions and lockdown, due to the high transmissibility of the virus(Sanche et al. 2020), many countries including Iran became afflicted within the first months of the outbreak(Hamzelou 2020).

Iran is one of the first countries to confront the Coronavirus outbreak with a high number of confirmed cases in the world. The government decided to close public places and cancel all social events to mitigate the transmission. By early March all provinces were affected by the pandemic which was just a few weeks after the first cases were reported in Qom (Venkatesan 2020). One great adversity faced during the pandemic was that the outbreak coincided with the Nowruz holidays which led to a sharp increase in number of daily cases (Heidari and Sayfouri 2020). On March 27^th^ in the middle of Nowruz vacations, Iranian authorities enacted travel restrictions to reduce the transportation of COVID-19 carriers to each province. the delay in decision making has been debated to be one of many reasons the disease spread rapidly across all provinces(Abdi 2020)

Many factors have been known to be associated with the initial levels of the Coronavirus outbreak at the country level including geographical factors(Sun et al. 2020), demographical parameters (Dowd et al. 2020), healthcare services(Emanuel et al. 2020), and economic status (Lai et al. 2020). Meanwhile, some research projects have been carried out to investigate the early-stage dynamics of the COVID-19 epidemic in Iran. For instance, (Dadar et al. 2020) convey that there is a negative relationship between COVID-19 incidence and the relative distance of provinces from Qom as the epicenter of the disease. Also, the study reported a relationship between the elderly population with the outbreak. In another study, (Ahmadi et al. 2020) reported a direct relationship between population density and intra-provincial movement with the infection rate of each province. This work also points out the weather impacts on the disease infection rate. Acknowledging such factors would be of great help in foreseeing inevitable conditions brought by epidemics in different climate zones.

Artificial Neural Network as a machine learning tool has been wildly used in studies to forecast the outbreak due to its proven application in epidemiologic risk assessments(Manliura Datilo, Ismail, and Dare 2019). (Car et al. 2020) has used a limited time-series dataset of Coronavirus patients to obtain a quality neural network model of disease spread. In another case in which artificial neural networks have become handy (Mollalo, Rivera, and Vahedi 2020), they collected a wide range of relevant factors and employed a neural network model to forecast COVID-19 incidence across the United States.

Interestingly most of the research studies in Iran were conducted during the period before travel restrictions in which It was permissible for provinces to receive travelers from other provinces. In this paper, we aim to generate a spread model of COVID-19 and investigate the demographical and healthcare factors impact on disease incidence from April 4^th^ to May 2^nd^ in 25 provinces. To predict daily confirmed cases in the provinces, we collected potentially impacting variables and employed a multi-layer perceptron network trained by publicly available COVID-19 data from Iran authorities. This work would give a vision of the dynamic nature of this epidemic in the early stages. Thus, both adopting policies and assessing mitigation strategies are crucial in the future.

## materials and Methods

### data collection

The number of daily confirmed cases for the period from April 4, 2020, to May 2, 2020, was obtained from the Iranian Ministry of Health and Medical Education website. The dataset shows the disease spread in provinces as time-series data. Due to the excess of missing values in reported cases, seven provinces are excluded from the study. Demographic data and literacy for each province were extracted from census data from the Statistical Centre of Iran (www.amar.org.ir) Table 2. To avoid any underlying association of variables, men’s population divided by province populations was used. Since each province has medical infrastructures to cope with COVID-19, Province-level healthcare infrastructure indices were extracted from (Shojaei et al. 2020) to represent healthcare access and quality of each province. Incidence rate per 100,000 residents during the study period (2 April until 4 May) with all variables is reported for provinces. The geographical distribution of the confirmed cases and healthcare indices are illustrated in **Error! Reference source not found.**.

**Table 2.** The descriptive statistics of the variables.

| variable | Var | Min | Max | Mean |
| --- | --- | --- | --- | --- |
| Distance from Qom | 114919 | 0 | 1344 | 648.917 |
| Population | 3E+09 | 11240 | 183285 | 56675 |
| Gender ratio (%) | 7E+12 | 580158 | 1E+07 | 2592877 |
| Distribution (%) | 3E-05 | 0.50242 | 0.5336 | 0.50846 |
| Over 65 years old (%) | 8.7169 | 58.5 | 73 | 69.125 |
| Urban population (%) | 0.0138 | 0.48491 | 0.9518 | 0.69049 |
| Density (population per area) | 34453 | 5 | 969 | 91.7083 |
| Literacy rate (%) | 12.582 | 76 | 92.9 | 85.825 |
| Health infrastructure quality | 0.0491 | 0 | 0.9879 | 0.78934 |

### Statistical analysis

Data preprocessing is necessary to reduce the effect of outliers in analyses. All the variables including population-based, health indices, literacy indices, altitude, and distance from the epicenter, were normalized before the analysis. Any linear association between incidence rate per 100,000 residents and the variables was measured via Pearson’s correlation coefficient (r). All analyses were performed using python version 3.8

### Artificial neural network

Multi-layer perceptron (MLP) is a common network model for supervised learning regression studies. It consists of input, hidden and output layers figure 1. In this study, the capability of each variable as predictors is evaluated through the network model to predict new COVID-19 cases of Iran provinces.

**Fig. 1.**
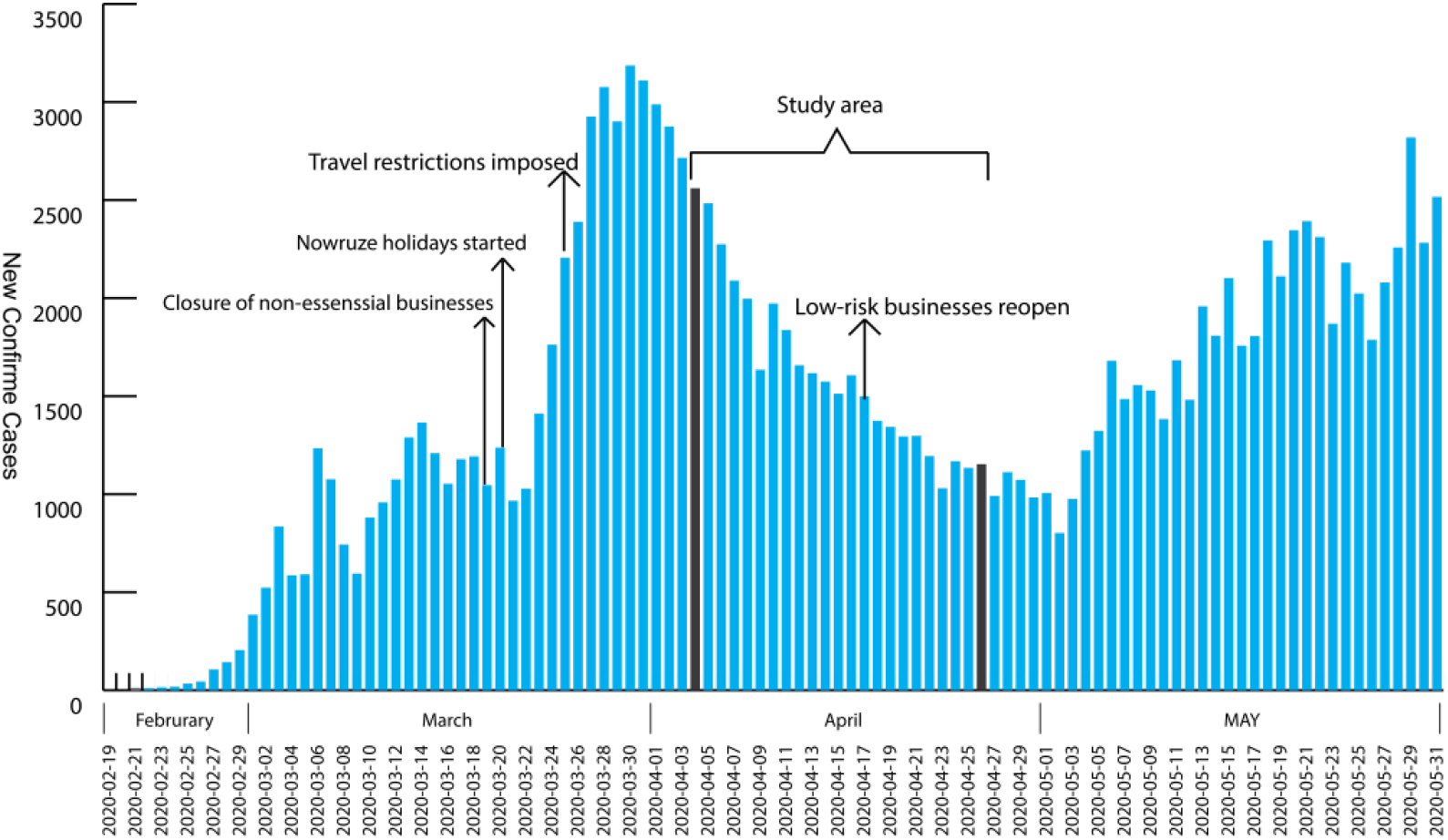
demographics of COVID-19 confirmed cases during early days of the pandemic in Iran.

The variables table 1 was imported as neurons into the network via the input layer. These values then were affected by weights of the connections and imported to the next hidden layers which ended to produce the output variable in the output layer. The flexible ability of hidden layers to recognize patterns in the data makes this machine-learning method superior to the standard regression model.

**Table 1.** Characteristics of provinces with reported COVID-19 cases used in the study.

| Province | Distance from Qom (km) | Population | Gender ratio (%) | Distribution (%) | Urban population (%) | Density (population per km<sup>2</sup>) | Literacy rate (%) | Health infrastructure quality index | Over 65 years old (%) | Incidence per 100,000 inhabitants |
| --- | --- | --- | --- | --- | --- | --- | --- | --- | --- | --- |
| Ardabil | 623 | 1270420 | 0.51 | 1.59 | 0.68 | 71.00 | 83.10 | 0.9271 | 6.4 | 231.10 |
| Bushehr | 897 | 1163400 | 0.53 | 1.46 | 0.72 | 51.00 | 89.20 | 0.9464 | 4.3 | 15.37 |
| Chaharmahal and Bakhtiari | 384 | 947763 | 0.51 | 1.19 | 0.64 | 58.00 | 84.70 | 0.9254 | 5.8 | 27.68 |
| East Azerbaijan | 657 | 3909652 | 0.51 | 4.89 | 0.72 | 86.00 | 84.70 | 0.7342 | 7.2 | 151.49 |
| Fars | 760 | 4851274 | 0.51 | 6.07 | 0.70 | 40.00 | 88.80 | 0.4157 | 6.1 | 50.70 |
| Hormozgān | 1141 | 1776415 | 0.51 | 2.22 | 0.55 | 25.00 | 87.80 | 0.8945 | 3.9 | 37.70 |
| Ilam | 596 | 580158 | 0.51 | 0.73 | 0.68 | 29.00 | 84.90 | 0.9196 | 5.6 | 36.68 |
| Isfahan | 281 | 5120850 | 0.51 | 6.41 | 0.88 | 48.00 | 89.90 | 0.4977 | 7.1 | 66.46 |
| Kerman | 847 | 3164718 | 0.51 | 3.96 | 0.59 | 17.00 | 85.40 | 0.7310 | 5.1 | 16.04 |
| Kermanshah | 425 | 1952434 | 0.51 | 2.44 | 0.75 | 78.00 | 84.50 | 0.8007 | 6.5 | 65.41 |
| Khūzestān | 666 | 4710509 | 0.51 | 5.89 | 0.75 | 74.00 | 86.30 | 0.6523 | 4.5 | 39.87 |
| Kohgiluyeh and Boyer-Ahmad | 611 | 713052 | 0.51 | 0.89 | 0.56 | 46.00 | 84.40 | 0.9879 | 4.6 | 38.91 |
| Kurdistan | 1154 | 1603011 | 0.51 | 2.01 | 0.71 | 55.00 | 81.50 | 0.9523 | 6.1 | 51.62 |
| Lorestan | 337 | 1760649 | 0.51 | 2.20 | 0.64 | 62.00 | 83.00 | 0.8723 | 5.8 | 89.07 |
| Markazi | 133 | 1429475 | 0.51 | 1.79 | 0.77 | 49.00 | 87.00 | 0.8642 | 7.6 | 35.85 |
| Mazandaran | 389 | 3283582 | 0.50 | 4.11 | 0.58 | 138.00 | 88.70 | 0.6364 | 7.6 | 81.64 |
| North Khorasan | 827 | 863092 | 0.50 | 1.08 | 0.56 | 30.00 | 83.30 | 0.9814 | 5.5 | 74.89 |
| Qom | 0 | 1292283 | 0.51 | 1.62 | 0.95 | 112.00 | 88.70 | 0.9465 | 4.8 | 137.22 |
| Semnan | 283 | 702360 | 0.51 | 0.88 | 0.80 | 7.00 | 91.50 | 0.8556 | 6.7 | 39.87 |
| Sistan and Baluchestan | 1344 | 2775014 | 0.51 | 3.47 | 0.48 | 15.00 | 76.00 | 0.7228 | 3.2 | 25.02 |
| South Khorasan | 1027 | 768898 | 0.51 | 0.96 | 0.59 | 5.00 | 86.70 | 0.9480 | 6.9 | 30.66 |
| Tehran | 1027 | 13267370 | 0.50 | 16.60 | 0.94 | 969.00 | 92.90 | 0.0000 | 6.8 | 63.68 |
| West Azerbaijan | 796 | 3265219 | 0.51 | 4.09 | 0.65 | 87.00 | 82.00 | 0.8106 | 5.6 | 61.35 |
| Zanjan | 369 | 1057461 | 0.51 | 1.32 | 0.67 | 49.00 | 84.80 | 0.9215 | 6.7 | 36.53 |

To restructure the time-series dataset of confirmed cases to a supervised learning problem, we used the sliding window method. An approximated window iteratively segments the time-series data until optimal window size(m) with the least possible error is chosen. A matrix of historical daily case data in all provinces is then retrieved to train the neural network. The output variable, in this case, is province-level forecasted daily COVID-19 cases.

*k-fold cross-validation* technique carried out for model internal validity assessment. This technique randomly partitioned the dataset to k subsets of equal size (in this case, *k*=10), which was repeatedly used in training and testing to the point every subset served once to test the model. The entire process was repeated 10 times. The result is then presented as the average of achieved scores, with standard deviation noted.

A base model which only contains daily cases as input variables were defined to assess the performances of neural network each time variables were added as an input to the model figure 2. Root mean square error (RMSE) and the absolute fraction of variances (R2) metrics were utilized to evaluate the performance of the model and to investigate the capabilities of variables to predict the output variables. All the procedure was performed in the python environment using scikitlearn library on a PC with a six-core 2.6 GHz processor and 16 GBs RAM.

**Fig. 2.**
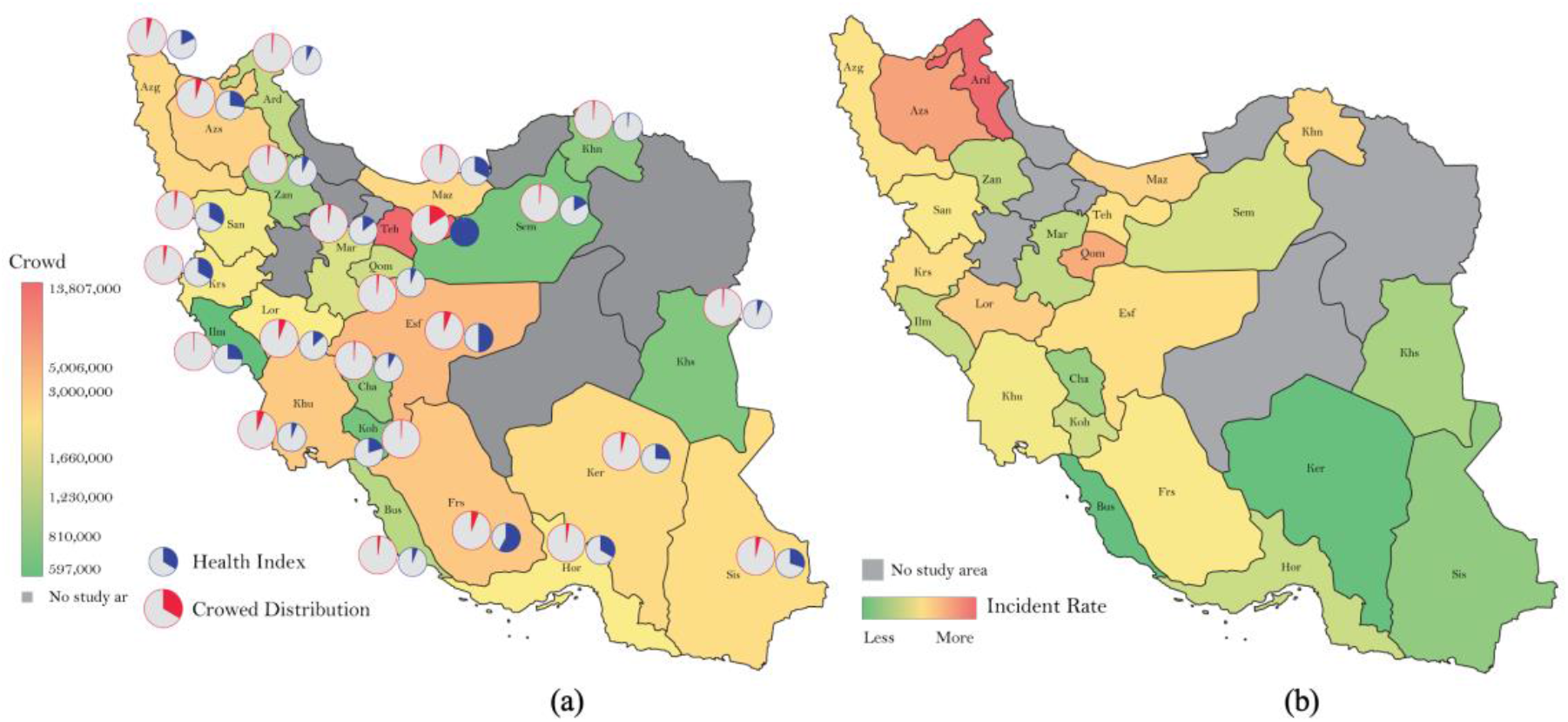
(a): Distribution of health infrastructure indices and population density in Iran, (b): COVID-19 incidence distribution across provinces in the study period.

## Results

Iran has a population of over 83,000,000 with a territory of 1,648,195 km^2^. All analyses were carried out in the period of 4 April until 2 May for 24 provinces. Most cases were reported in Tehran as a highly-populated and developed province.

The linear relationships between COVID-19 incidence and variables were investigated Table 3. The results revealed that COVID-19 incidence in provinces has no significant associations with either demographic variables or healthcare indices. Also, no statistically significant relation was found between disease incidence in provinces with their Distance from Qom as the epicenter of the disease (r = −0.26, p > .05).

**Table 3.** Correlation analysis between Correlation between the incidence of COVID-19 and the variables.

| Variable | Pearson |  | Spearman |  |
| --- | --- | --- | --- | --- |
|  | Correlation | *p*-value | Correlation | *p*-value |
| Distance from Qom (km) | -0.26 | 0.21 | -0.31 | 0.14 |
| Population | 0.03 | 0.87 | 0.32 | 0.13 |
| Over 65 years old population (%) | 0.26 | 0.22 | 0.34 | 0.1 |
| Urban population ratio (%) | 0.26 | 0.21 | 0.31 | 0.14 |
| Literacy rate (%) | -0.06 | 0.79 | -0.1 | 0.64 |
| Population distribution (%) | 0.04 | 0.85 | 0.32 | 0.13 |
| Health infrastructure quality | 0.02 | 0.92 | -0.11 | 0.62 |
| Gender ratio (%) | -0.08 | 0.69 | -0.15 | 0.47 |
| Population density (population per area) | 0.09 | 0.66 | 0.61 | 0.001 |

A multilayer perceptron was used in the study. The optimal number of hidden layer neurons was obtained by a trial-and-error procedure and selection was made based on the lowest RMSE and highest R2 metrics. In the MLP model, 18 combinations of inputs were examined to evaluate the performance of variables to act as predictors, each time they were added as an input to the base model figure 2. Most accurate models were obtained when variables population distribution(R2=84%), health index(R2=82%), and population(R2=86%) were added as an input to the base model Fig 4.

**Fig. 3.**
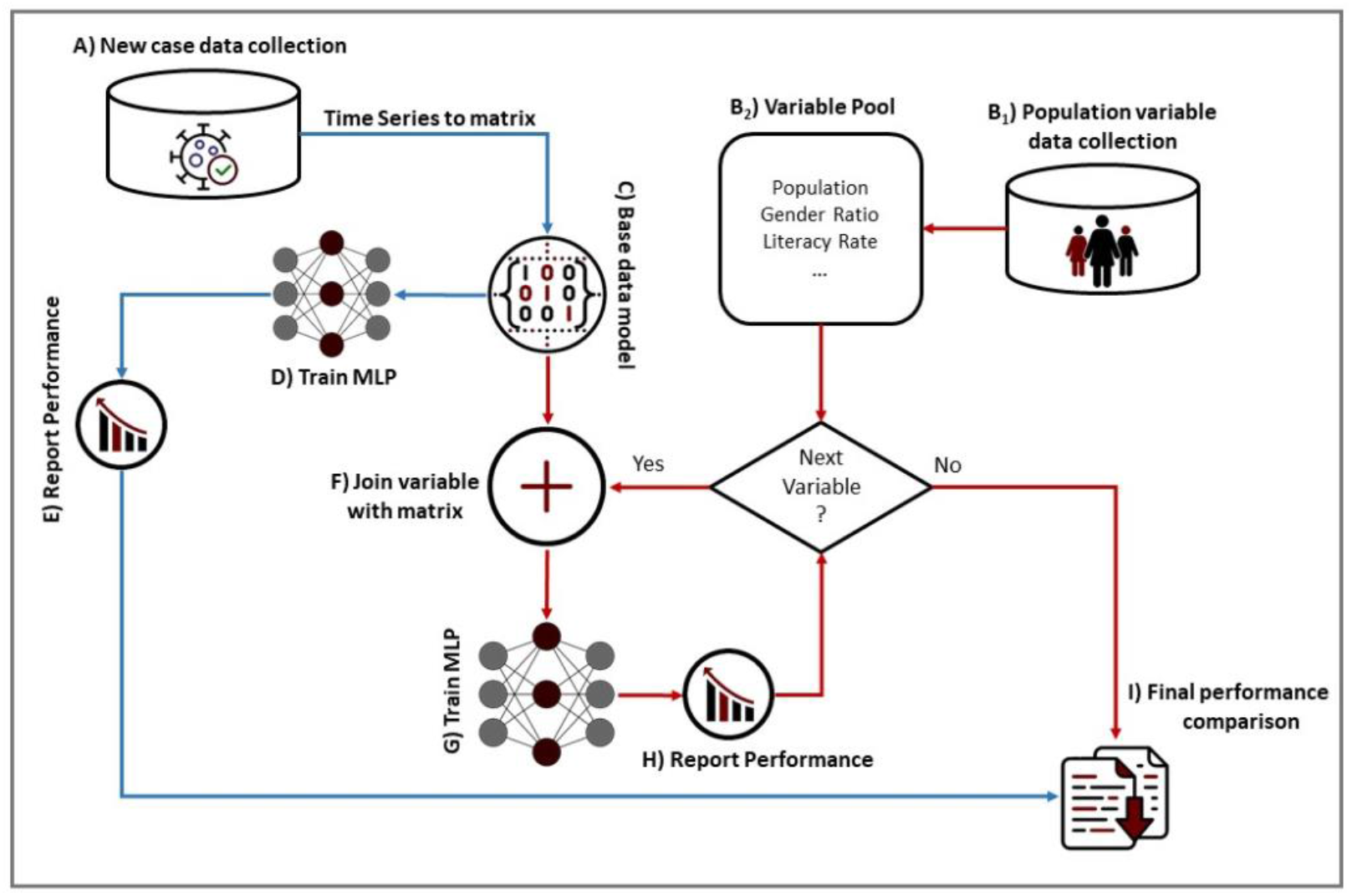
Flow diagram of modeling process. Confirmed cases data is collected (A) and converted to a base matrix, the matrix is then used to train the MLP(D) with a performance report (E). Each demographic variable(B_1_) is added to base matrix in a loop like manner(F) to train the network(G) with a performance report(H), the performances are compared to investigate the effects.

**Fig. 4.**
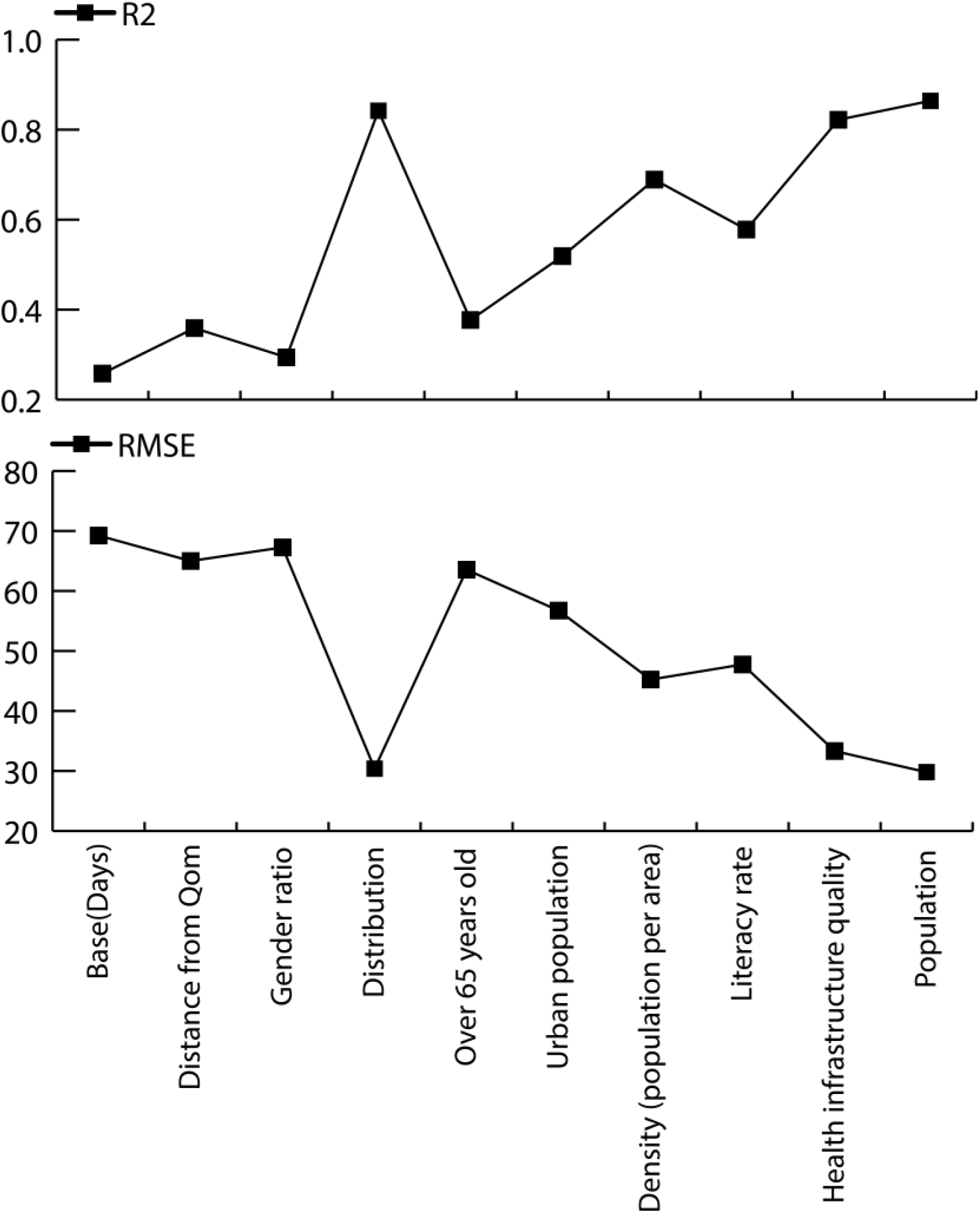
Summary comparative performance reports of the models.

## Discussion

Coronavirus’s early-stage spread was investigated in this study with the help of an artificial neural network due to its proven potentials in epidemic forecasting. New Covid-19 cases were predicted with the use of daily diagnosed cases from 25 provinces of Iran from 2 April until 4 May, along with demographic and healthcare factors to discover new possible influencers of the disease spread in the country.

The most notable variable is the distance from Qom which shows how provinces are vulnerable to spread from the epicenter of the disease. It was shown in a study(Dadar et al. 2020) in the earlier days of the outbreak, the distance was significantly associated with COVID-19 spread. Our study showed that no significant relationships existed in the observation period of the study. One worth-mentioning issue is that the Nowruz holidays had a great impact on the spread dynamics of the pandemic(Heidari and Sayfouri 2020). Our study was carried out in the period after the holidays in which many travelers had contributed to rapid growth in daily COVID-19 cases and disease spread across the country. This can be in tune with the postulation that with the amount of time after all provinces are affected by the virus, especially after holidays, the distance would not be a criterion for predicting the epidemic.

The study aimed to develop a COVID-19 spread forecasting model in search of variables’ contribution to disease prevalence. Models with high accuracy were obtained by taking population, population density, and health indices to account as predictor variables in the neural network. Thus, no direct relationships were found between population density or population with the incidence rate of COVID-19, both factors were effectual in the increase of accuracy of our predictive model. Also, Population density had been proved to be an effective parameter in the outbreak as mentioned in (Ahmadi et al. 2020) in earlier days of the pandemic which could be a sign of dynamic behavior of the pandemic.

In the course of our research, we faced some limitations mostly due to the lack of data in the case of both variables and daily cases. Many factors may be involved in the virus spread across the country which was not accessible at the province level. Also, we were not able to verify our predicted data to the real data since the data was not published for provinces.

characterizing the main factors of the early-stage outbreak of such a contagious virus provides opportunities to comprehend the early dynamics of the outbreak. Also, it could be of great importance in confronting the future epidemics, in terms of taking preventive action or implementing control measures by policymakers based on prior knowledge.

## Data Availability

All variable data for each province were extracted from census data from the Statistical Centre of Iran and conformed cased from the Iranian Ministry of Health and Medical Education website.

